# COVID in Post Vaccinated individuals- Beacon of Light

**DOI:** 10.1101/2021.06.18.21258796

**Authors:** VK Sashindran, K Rajesh, Ankur Nigam, Sourya Sourabh Mohakuda, Navin Bhati, D Mohapatra

**Affiliations:** Dept of Internal Medicine, 7 Air Force Hospital, Kanpur; Public Health Specialist, Gangtok

**Keywords:** COVISHIELD, COVAXIN, EFFICACY, VACCINATION

## Abstract

**Introduction:** COVISHIELD and COVAXIN have been introduced post rapid approval as COVID vaccines in India, which has the second most COVID cases across countries. These vaccines are being administered in a two-dose schedule from 16 Jan 2021. This study deals with the clinical profile of individuals who developed COVID infection post-COVID vaccination.

This is the first study of similar nature in India.

**Methodology:** The study population comprised of individuals who were detected to be COVID positive 04 weeks post-vaccination and were compared with individuals detected positive within the first 04 weeks of vaccination. Data was collected in a digital questionnaire format and analyzed with SPSS v-23 software. Clinical features were profiled in detail. Chi-square analysis was done to find out the association of various demographic features with the severity of the disease.

**Results:** In the study population, fever was the commonest symptom (75.1%) followed by anosmia (72.1%), and shortness of breath (16.3%). There was a lower incidence of fever, cough, dyspnea, and requirement of hospitalization in the study population as compared to the control group and previous epidemiological data. The time required for complete recovery and disease severity was favorable in our study population. There was a significant correlation in the rate of hospitalization among the study group and the comparative group (p=0.0001) and between the number of dosage of COVID vaccine with the lowest SpO2 recorded (p=0.001).

**Conclusion:** This study will boost the ongoing initiative of having a maximal vaccinated population countrywide and emphasize the need for two doses of vaccination.

## Introduction

Severe Acute Respiratory Syndrome Corona Virus 2 (SARS CoV2), a coronavirus, causing COVID 19 infection was declared a pandemic on 11 Mar 2020 by the World Health Organization [1]. The pandemic has not only inflicted mortality and morbidity across the human race but has also crippled the economies of affected nations. The imperative to control this disease has encouraged vaccine research worldwide. The severe second and third waves of infections have made the world realize that rapid vaccination of as many people as possible with an effective vaccine is the only lasting solution.

Vaccination against the disease was launched in India on 16 Jan 2021 with healthcare workers, frontline workers, and elderly above 60 years being the first beneficiaries. Two vaccines have so far been used in India: COVISHIELD and COVAXIN. The COVISHIELD vaccine is a viral-vectored vaccine, developed by the Serum Institute of India in line with the vaccine developed in the UK by Jenner Institute, University of Oxford. COVAXIN is an inactivated virus vaccine, developed by Bharat Biotech in collaboration with the Indian Council of Medical Research [2].

During the early stages of vaccination, there were public concerns about the safety and efficacy of the vaccines. This was mainly due to the rapid approvals given to their usage [3, 4]. Subsequently, several scientific publications have validated the safety of these vaccines [5]. The efficacy as determined by seroconversion after the first and second doses of COVISHIELD is 91% and 100% respectively and that of COVAXIN is 98.3% at 56 days post-vaccination [6,7].

Various studies have also shown that vaccination confers immunity and prevents progression to severe disease. In this study, we have highlighted the clinical profile of patients who were detected to be COVID positive post-vaccination. To the best of our knowledge, this is the first paper on the analysis of the clinical profile of COVID illness in post-vaccination individuals in India.

## Materials and methods

This descriptive observational study was conducted in individuals under the area of responsibility of a tertiary care center in Northern India with a premise to study the clinical profile of patients with COVID infection and various factors associated with severity of disease post-COVID vaccination. All COVID positive patients who developed COVID at least 4 weeks after the first dose of COVID vaccination were eligible for enrollment. Informed consent was taken from all participants. The subjects were administered a questionnaire via digital means during the period from Apr 2021toMay 2021. Data of all the beneficiaries were compiled and collated in an excel sheet and was analyzed after data cleaning using SPSS v-23.

This is the first study highlighting clinical features in COVID positive individuals post-vaccination to the best of our knowledge. Individuals who were tested positive by rapid antigen test/RTPCR were taken as COVID positive patients. The lowest oxygen saturation on pulse oximeter (Spo2) recorded by the study participants was used as a surrogate marker to assess the severity of the disease. The mild disease was said to be present if the lowest Spo2 recorded by study participant was more than or equal to 94%, SpO2 of 90%-93%, and below 90% were considered moderate and severe COVID respectively. For this study, post-vaccination COVID infection was defined as a confirmed COVID positive case who has taken at least one dose of COVISHIELD and COVAXIN four weeks before having COVID infection. This time cut-off was taken as the time taken for seroconversion after COVID vaccination is approximately 04 weeks [6,7]. Those who developed COVID less than four weeks after the first dose of vaccination were enrolled in the comparative arm of the study.

### Statistical analysis

Descriptive statistics (frequency and percentages) were calculated for sample demographic characteristics and clinical features of the participants. Chi-square analysis was done to find out the association of various demographic features with the severity of disease among the study participants. The data of individuals with COVID infection following COVID vaccination after single and two doses was analyzed separately and interpreted. Binary logistic regression was done for variables that were found to have a statistically significant association with the outcome of interest.

## Results

There were 168 participants in the study group and 42 participants in the comparative group whose mean age was 37.18(+/- 9.38) & 40.36(+/- 11.274) respectively. The study population comprised 140 (83.3%) males and 28 (16.7%) females in the study group & 31 (73.8%) males and 11(26.2%) females in the comparative group respectively. The study population was divided into 05 subgroups based on their age profile. 33(19.6%) of the participants had at least one comorbid condition against 16 (38.1%) in the comparative group. Demographic features of the entire study population are given in Table-01.

**Table-01:** Demographic features of the study population & comparative group.

|  | **Study population**<br>**Total (N=168) n(%)** | **Comparative group**<br>**(N=42) n(%)** |
| --- | --- | --- |
| **Gender** |  |  |
| Male | 140 (83.3) | 31 (73.8) |
| Female | 28 (16.7) | 11 (26.2) |
| **Age (years)** |  |  |
| 20-30 | 40 (23.8) | 12 (28.6) |
| 30-40 | 73 (43.5) | 12 (28.6) |
| 40-50 | 34 (20.2) | 09 (21.4) |
| 50-60 | 19 (11.3) | 08 (19.0) |
| 60-70 | 02 (1.2) | 01 (2.4) |
| **Comorbidity** |  |  |
| Present | 33 (19.6) | 16 (38.1) |
| Absent | 135 (80.4) | 26 (61.9) |
| **Profession** |  |  |
| Health care worker | 54 (32.1) | 02 (4.8) |
| Front line worker | 64 (38.1) | 12 (28.6) |
| Others | 50 (29.8) | 28 (66.7) |

**Table-02:** Vaccination details of the study population.

|  |  |  |
| --- | --- | --- |
|  | **Total (N=168) n(%)** |  |
| **Type of Vaccine administered** |  |  |
| Covishield | 162 (96.4) |  |
| Covaxin | 06 (3.6) |  |
| **Number of doses administered** |  |  |
| First dose | 10 (6.0) |  |
| Second dose | 158 (94.0) |  |
| **Duration after vaccination when the individual became COVID positive** |  |  |
|  | **After 1<sup>st</sup> Dose** | **After 2<sup>nd</sup> Dose** |
| Less than 2 weeks | - | 25 (15.8) |
| 2-4 weeks | - | 34 (21.5) |
| 4-8 weeks | 10 (100) | 74 (46.8) |
| >8 weeks | - | 25 (15.8) |

One hundred and twenty-nine (76.8%) of the study participants were symptomatic for COVID infection and 29(17.3%) of them required hospitalization. Twenty-nine (69.0%) of the comparative group were symptomatic for COVID infection and 20 (47.6%) of them required hospitalization. Fever was the predominant symptom 122 (77.2%), followed by loss of smell/taste and cough among the study group participants. A similar trend was observed among the comparative group. A significant association was found for shortness of breath and the lowest SpO2 recorded among the participants when compared in both arms. Clinical features of the study population and comparative group with symptomatic COVID infection are given in Table-03.

**Table-03:** Clinical features of the study population & comparative group among symptomatic individuals.

| Clinical Characteristics |  | Study population<br>Total n (%) | Comparative group n<br>(%) | p-value |
| --- | --- | --- | --- | --- |
| **1. Fever** |  |  |  |  |
| (a) Present |  | 97 (75.1) | 25 (86.2) | .201 |
| Absent |  | 32 (24.9) | 04 (13.8) |  |
| (b) Duration of fever | <5 days | 69 (71.1) | 17 (68.0) |  |
|  | 5-10 days | 25 (25.8) | 08 (32.0) |  |
|  | >10 Days | 03 (3.1) | - |  |
| (c) Maximum<br>recorded temp | <100 | 27 (27.8) | 08 (32.0) |  |
|  | 100-102 | 64 (66.0) | 13 (52.0) |  |
|  | 102-104 | 05 (5.2) | 03 (12.0) |  |
|  | >104 | 01 (1.0) | 01 (4.0) |  |
| **2. Cough** |  |  |  |  |
| (a) Present |  | 90 (69.8) | 19 (65.5) | .654 |
| Absent |  | 39 (30.2) | 10 (34.5) |  |
| (b) Type of cough | Dry | 65 (72.2) | 11 (57.9) |  |
|  | Productive | 25 (27.8) | 08 (42.1) |  |
| (d) Duration of cough | <5 days | 32 (35.6) | 07 (36.8) |  |
|  | 5-10 days | 28 (31.1) | 10 (52.7) |  |
|  | >10 Days | 30 (33.3) | 02 (10.5) |  |
| **3. Shortness of breath** |  |  |  |  |
| (a) Present |  | 21 (16.3) | 08 (27.6) | .004 |
| Absent |  | 108 (83.7) | 21 (72.4) |  |
| (b) Onset of shortness of breath | <2days | 08 (38.1) | 02 (25.0) |  |
|  | 2-6 days | 10 (47.6) | 04 (50.0) |  |
|  | >6 days | 3. (14.3) | 2. (25.0) |  |
| **4 Lowest Spo2 recorded** | 80-90 | 02 (1.6) | 03 (10.3) | .001 |
|  | 90-94 | 13 (10.0) | 08 (27.6) |  |
|  | >94 | 114 (88.4) | 18 (62.1) |  |
| **5 Loss of smell/taste** |  |  |  |  |
| (a) Present |  | 93. (72.1) | 22. (75.9) | 0.169 |
| Absent |  | 36 ( 27.9) | 07 (24.1) |  |
| **6. Gastrointestinal symptoms** |  |  |  |  |
| (a) Present |  | 42 (32.6) | 09 (31.0) | 0.874 |
| Absent |  | 87 (67.4) | 20 (69.0) |  |

The Chi-square test was used to find out the association of various demographic features with the outcome of interest among the study participants. It was found that the number of dosage of the COVID vaccine had a significant association with the lowest Spo2 recorded among the study participants (Table-04).

**Table-04:**
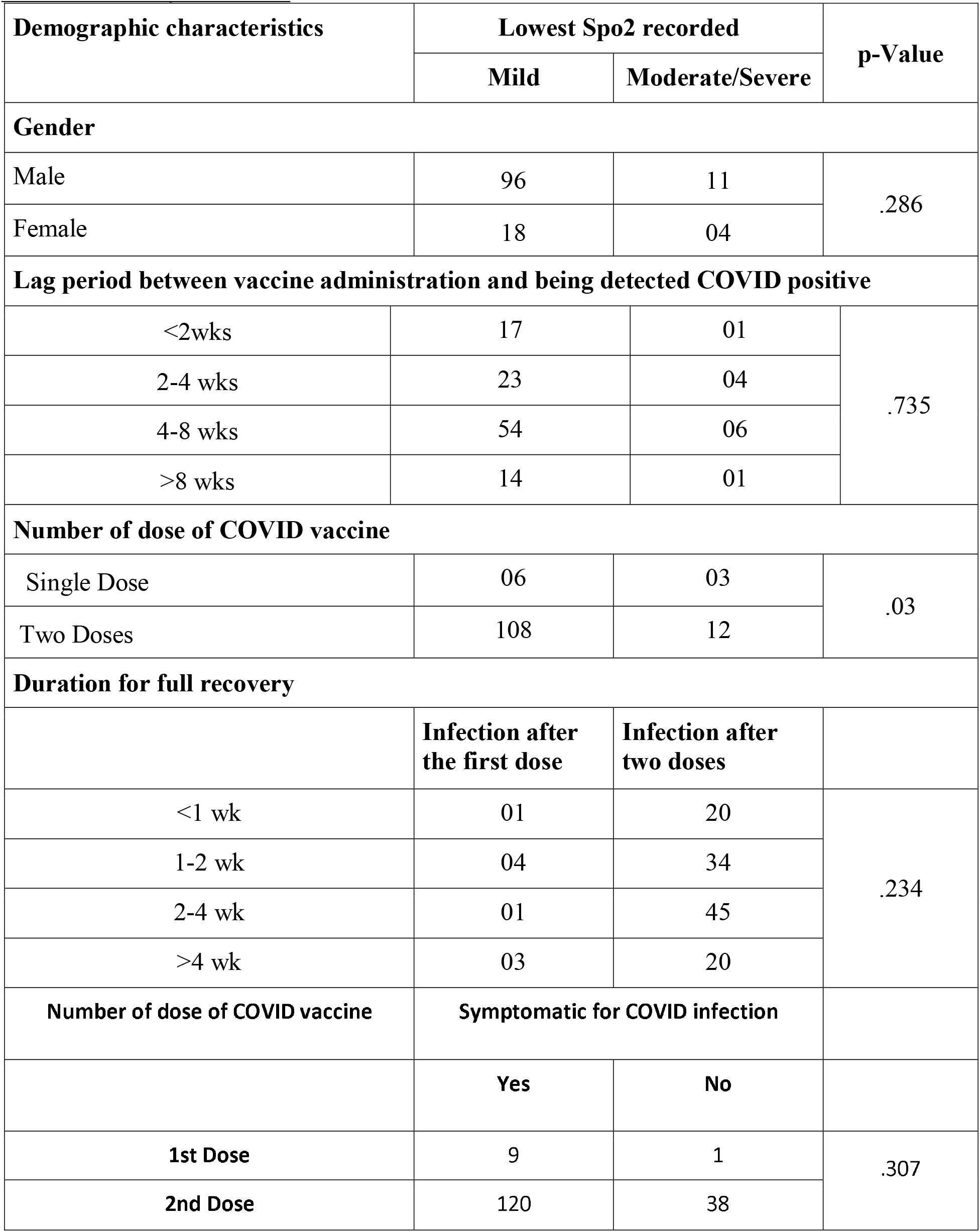
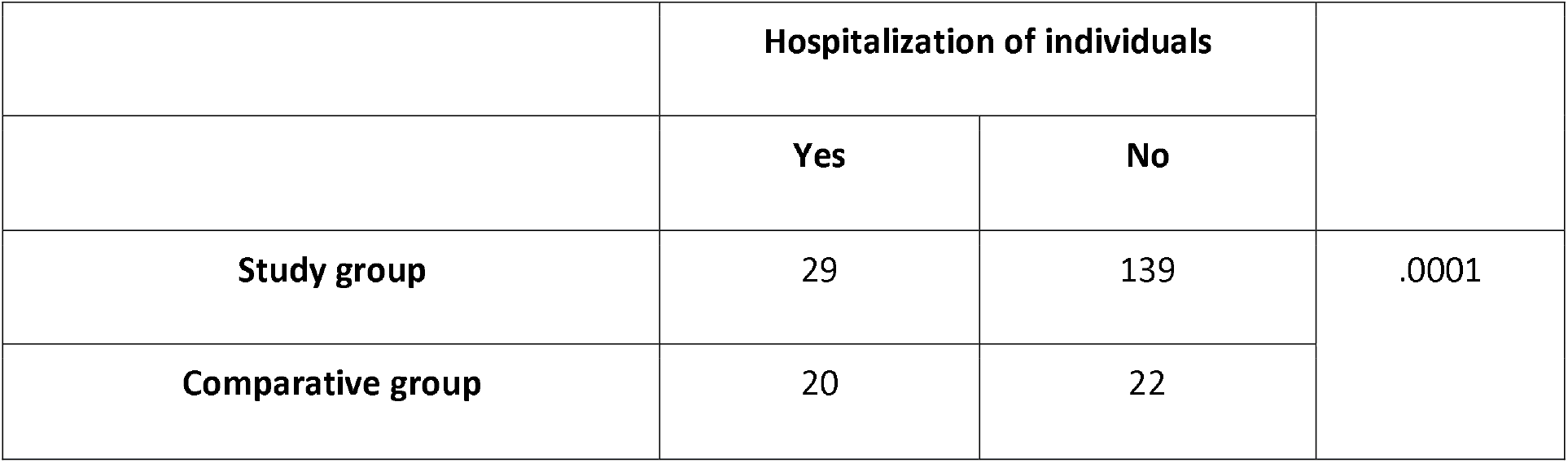
Association of various demographic characteristics among the study participants with the severity of disease.

Binary logistic regression was further applied to the variables which had a significant association with the outcome of interest. The odds of milder infection after both the doses of COVID vaccine among the study group was 1.5 times that after a single dose (p=.05) (Table-05).

**Table-05:** Binary Logistic regression for COVID Vaccine minor reactions by demographic characteristics.

|  | OR | SE | 95% CI | p-Value |
| --- | --- | --- | --- | --- |
| **Mild Infection after the first dose** | REF | REF | REF |  |
| **Mild Infection after two doses** | 1.5 | .770 | 0.995-20.34 | .05 |

## Discussion

Initial epidemiological data shows that COVID-19 disease presents as mild cases in 81%, severe cases in 14%, and as critical cases in 5%. It is also associated with a mortality rate of about 2.5% [8]. In addition, clustering of cases in a limited geographical area over a short time overwhelms health care facilities. Mass vaccination is considered the only viable strategy to mitigate viral pandemics. This descriptive study demonstrates the clinical profile of individuals developing COVID infection post-vaccination.

Our study population was relatively young with a mean age of 37.18 (+/-9.38) and this was comparable with the comparative population. The proportion of individuals with co-morbid conditions is comparable to the general population [9]. With this second wave of COVID affecting the younger population, our study population can be considered as a suitable representation of the general population [10]. Most of our study population (96.4%) received COVISHIELD with 94% of the study population having taken both doses of vaccination. As the subjects did not have a choice as to which vaccine they could take, interpretation on the choice of the vaccine was not done.

Fever was the most predominant symptom, followed by loss of smell/taste in 75.1% and 72.1% of the study population respectively. In a meta-analysis done by Li et al., fever was seen in 88.5%, cough in 68.6%, dyspnea in 21.9%, and gastrointestinal symptoms in 8.7% of symptomatic individuals [8]. There was a lower incidence of fever, cough, and shortness of breath in our study population as compared to both the comparative group and the study by Li et al. However, the above-referenced study was only a meta-analysis of the clinical profile of COVID patients and did not focus on post-vaccination COVID.

The clinical profile of post-vaccination COVID was that of a short febrile illness with 70.5% of the febrile patients having fever for less than 5 days. In a study on the clinical profile of COVID infection in the non-vaccinated population, Raman Sharma et al found the median duration of fever to be 12 days [11]. The duration of fever is considerably lower among our vaccinated subjects. As per our SpO2 based severity classification, 83.5% had mild, 13.3% moderate, and 3.2% severe disease. Saturation below 80% was not recorded in any of our subjects. In the study by Li et al, on non-vaccinated COVID patients, 14% had severe and 5% had critical disease[8]. Vaccination thus seems to mitigate critical and severe disease.

The duration for complete recovery in our study as shown in the results is considerably faster than that of data published by MP Barman et al where less than 4% of the patients had recovered in 10 days and it took 25 days for 50% of the patients to recover fully [12].

There was a significant association (p=0.0001) with a decreased rate of hospitalization among the study population as compared to the comparative group. This is much lower than the average rate of hospitalization required in COVID among unvaccinated individuals.[13]

There was a significant correlation between the number of dosages of the COVID vaccine with the lowest SpO2 recorded (p=0.001) with the odds of having mild infection after both doses of the vaccine among the study group was 1.5 times than that after a single dose. (p=0.05). Despite an extensive search, we could not find any published study analyzing similar data. This translates into a better outcome of COVID with two doses of vaccine.

## Conclusion

This study on post-vaccination COVID infection in India shows the numerous benefits of vaccination and should be a guiding light for the administration to continue striving to vaccinate most of the population quickly.

## Data Availability

the data has been collected by administering the study participants with a digitalized questionnaire.

